# Flushing of stagnant premise water systems after the COVID-19 shutdown can reduce infection risk by *Legionella* and *Mycobacterium* spp

**DOI:** 10.1101/2020.09.14.20194407

**Authors:** Raymond M. Hozalski, Timothy M. LaPara, Xiaotian Zhao, Taegyu Kim, Michael B. Waak, Tucker Burch, Michael McCarty

## Abstract

The unprecedented widespread closing of buildings due to the COVID-19 pandemic has allowed water to stagnate in premise plumbing systems, creating conditions that may facilitate the growth of opportunistic pathogens. In this study, we flushed and collected samples from showers in buildings that had been unoccupied for approximately two months and quantified *Legionella pneumophila* using a commercial cultivation-based assay. In addition, all bacteria, *Legionella* spp., *L. pneumophila, L. pneumophila* serogroup 1, non-tuberculous mycobacteria (NTM), and *Mycobacterium avium* complex (MAC) were analyzed using quantitative PCR (qPCR). Despite low or negligible total chlorine in the stagnant pre-flush water samples, *L. pneumophila* were not detected by either method; *Legionella* spp., NTM, and MAC, however, were widespread. Using quantitative microbial risk assessment (QMRA), estimated risks of clinical illness from exposure to legionella and MAC via showering were generally low, but the risk of subclinical infection via *Legionella* spp. could exceed a 10^-7^ daily risk threshold if just a small fraction (≥0.1 %) of those legionellae detected by qPCR are highly infectious. Flushing cold and hot water lines rapidly restored a total chlorine (as chloramine) residual and decreased all bacterial gene targets to building inlet water levels within 30 min. Following flushing, the chlorine residual rapidly dissipated and bacterial gene targets rebounded, approaching pre-flush concentrations after 6 to 7 days of stagnation. These results suggest that stagnant water in premise plumbing may contain elevated levels of opportunistic pathogens; flushing, however, can rapidly improve water quality and reduce the health risk but the improvement will be short-lived if building disuse persists.

## Introduction

The COVID-19 pandemic resulted in the abrupt closure of schools, gyms, restaurants, retail shops, offices, and other facilities, which remained empty except for perhaps maintenance or cleaning. Building inactivity results in stagnant water in the premise plumbing that can adversely affect water quality, including the loss of residual chlorine, growth of bacteria, and release of harmful metals like lead.^1-4^ Temporarily idle buildings is not a new issue (e.g., during the annual summer recess of schools). Yet, there are gaps in the peer-reviewed literature regarding microbiological water quality changes during such closures and the corresponding risks. Specifically, it is unclear how the unprecedented sudden temporary closure of buildings due to the recent pandemic will affect the prevalence of and potential human exposure to opportunistic pathogens of genera *Legionella* and *Mycobacterium*.

Legionellae and non-tuberculous mycobacteria (NTM) occur naturally in water and include clinically notable species, especially *Legionella pneumophila* (causative agent of Legionnaires’ disease and Pontiac fever) and *Mycobacterium avium* complex (MAC) (causative agent of pulmonary and other infections).^5^ Other species of either genera, however, may also cause disease, and underreporting and under-diagnosis are believed to be significant challenges globally.^6-8^

Legionellae have been observed in drinking water distribution networks and premise plumbing when there is little or no residual free chlorine (HOCl) but largely absent otherwise, especially when residual chloramine is maintained in the water (as monochloramine, NH_2_Cl).^9-16^ Conversely, NTM are known for their resistance to disinfectants and other antimicrobial agents, which enables their persistence in distribution networks containing chlorine or chloramine.^16-19^

One approach for addressing pathogen concerns in premise plumbing is flushing. Cold and hot water flushing is recommended at least twice weekly in U.S. hospitals to mitigate *L. pneumophila* in low-use or low-flow outlets.^20^ In chlorinated and chloraminated water systems, flushing can replenish disinfectant residuals and either decrease or increase water temperatures in the cold and hot water lines, respectively, to outside the optimal range for pathogen growth (about 25 to 42 °C).^5^

In this study, showers in five university buildings that had been wholly or largely unused for more than two months due to the COVID-19 pandemic were investigated prior to and during flushing to assess the stagnant water quality and the changes that occur during flushing. These buildings were located in two different cities, each supplied lime-softened, filtered, and chloraminated river water from separate treatment and distribution systems. Conventional water quality indicators (total chlorine, temperature, and pH) were measured onsite and microbiological water quality was assessed using culture- and DNA-based techniques. Additional samples were collected from these showers for up to a week after flushing to assess water quality changes during the post-flushing stagnation period. Finally, quantitative microbial risk assessment (QMRA) was performed to assess the health risks of showering with water that had stagnated for an extended period and the risk reduction benefits of flushing.

## Experimental Materials and Methods

### Site Selection

In response to the COVID-19 pandemic, the University of Minnesota, Twin Cities (UMN) suspended or significantly reduced most activities in March 2020. Five UMN buildings were selected for accessibility, their known inactivity during the pandemic (i.e., according to facilities management personnel and water meter readings), and to provide varying building ages (constructed from 1935 to 2014; Table S1). Four buildings (designated A to D) were located in Minneapolis, Minnesota (United States), and one was located in St. Paul, Minnesota (building E). The buildings comprised from 3 to 7 levels or floors and ranged in total gross area from 5300 to 28 700 m^2^. Two conventional showers—one each for hot and cold water—were arbitrarily selected and investigated per shower room out of approximately 8 to 32 showers per room. One shower room was investigated per building, except buildings D and E, where a secondary shower room was also analyzed. Fourteen showers total were sampled in seven shower rooms.

The Cities of Minneapolis and St. Paul are served by separate drinking water systems. Both systems withdraw water from the Mississippi River, perform lime softening, filtration, and disinfection, and then distribute the water with a chloramine residual of approximately 3.5 mg/L Cl_2_.

### Collecting Samples

Buildings were investigated during May and June, 2020 (Table S2). Before flushing showers, the quality of the municipal water supply was assessed by sampling as close as possible to the building entry point (“inlet”). The inlet was first flushed for 5 to 10min until steady-state was reached with respect to temperature and pH, after which total chlorine was measured and a 1 to 2 L sample was aseptically collected into either an autoclave-sterilized polypropylene bottle or a manufacturer-sterilized Whirl-Pak bag (Nasco; Fort Atkinson, Wisconsin). In the shower rooms, shower heads were removed and the shower neck was flame-sterilized. The cold or hot water was turned on and the first 1 to 2 L of water was collected and designated the “pre-flush” sample (time *t* = 0min). The showers remained on and additional water samples were collected at *t* = 6, 15, 30, and 45 min, for a total of 5 samples per shower. In all cases, the cold and hot water flushes were staggered for ease of sampling with the cold water flush initiated first and the hot water flush initiated 20 min later. Immediately upon sample collection, aliquots were removed for testing of total chlorine, temperature, and pH. The remainder was placed on ice for immediate transport to the laboratory. In buildings with a second shower room, the secondary showers were only flushed for 30 min and only two microbiological samples were collected (*t* = 0 and 30 min). Water samples were subsequently collected from every shower 2 to 4 days later (follow-up 1) and again 6 or 7 days later (follow-up 2) to assess post-flushing changes in water quality. Additional sample collection details are provided in the Supporting Information (SI).

### Sample Processing

Culturable *L. pneumophila* were enumerated by the Legiolert Quanti-Tray test (IDEXX Laboratories; Westbrook, Maine) using the manufacturer’s potable water protocol. Triplicate tests were performed on all but 10 samples (7 duplicates and 3 single tests) during a temporary shortage of supplies. Periodic testing of positive and negative controls were within manufacturer specifications. The bulk of the remaining water (590 to 998 mL) was vacuum-filtered to collect the microorganisms using a 47 mm mixed cellulose polymer membrane (0.2 μm pore size; MilliporeSigma; Burlington, Massachusetts). DNA was extracted and purified from filter membranes, as previously detailed.^14^ Negative controls were collected with each sample set (approximately 1 per 14 microbiological samples) by filtering 2 mL sterile water through a clean membrane.

### Chlorine decay experiment

The chloramine decay rate in Minneapolis tap water was experimentally determined. Triplicate glass bottles (1L) were baked at 550 °C for 6h to eliminate organic carbon and then filled with cold tap water (after flushing approximately 40 min). Total chlorine, pH, and temperature were periodically measured for 3 weeks. Chlorine decay rate constants were determined by nonlinear (weighted) least-squares estimates in an exponential decay model using function *nls()* in the R “stats” package.^21^

### Real-time qPCR

Marker genes were targeted by real-time quantitative polymerase chain reaction (qPCR) to quantify total *Bacteria* (16S rRNA genes^22^), genus *Legionella (ssrA*^23^), species *L. pneumophila (mip*^23^), *L. pneumophila* serogroup 1 (wzm^23^), environmental NTM (genus *Mycobacterium atpE*^24^), and MAC (16S-23S internal transcribed spacer (ITS) region^25^), using qPCR reaction chemistry and amplification protocols previously described.^14,18^ Additional information on the qPCR can be found in the SI.

### Data Analysis and Statistics

To determine the effects of flushing and the post-flushing stagnation period, marker gene concentrations were compared using either paired maximum likelihood estimation (MLE) with survival models (“survival” package) or Fong’s modified sign test via R software.^21,26-29^ Marker gene concentrations were also assessed for correlation with water quality parameters (total chlorine, temperature, and pH) using a multivariate approach—supplemental fitting of the parameters after principal components analysis (PCA) (“vegan” package) of the marker gene concentrations as ranks.^26,30^ Spearman’s rank correlations were utilized as alternative. Statistical methods and treatments are thoroughly detailed in the SI.

### Quantitative Microbial Risk Assessment

Risks of illness from exposure to *Legionella* spp. and MAC while showering were quantified via QMRA, using flushing period gene marker concentrations and the general approach of Hamilton et al.^31,32^ Relevant health endpoints for legionellae were subclinical respiratory infection (i.e., Pontiac fever) and clinical severity respiratory infection (i.e., Legionnaires’ disease),^32^ with transmission modeled via aerosol inhalation during showering. Relevant health endpoints for MAC were respiratory infection (via aerosol inhalation while showering), systemic (disseminated) infection, and cervical lymphadenitis.^31^ The latter two are non-respiratory infections with transmission modeled via accidental ingestion of water during showering.

Inhaled or ingested doses were calculated using equations from Hamilton et al.,^32^ with input values from Ahmed et al.^33^ and a dose harmonization factor added for conversion of marker genes to colony-forming units (CFU), as previously used by Lee et al.^11^ and Ditommaso et al.^34^ Inhaled doses were determined for conventional and high-efficiency shower heads (13 L/min and 7 L/min, respectively). More details are provided in the SI.

Water concentrations depended on availability of non-zero observations for the pathogen of interest. For *Legionella* spp. in both cities and MAC in Minneapolis, concentrations were either the observed values of *ssrA* and ITS or 0 (for non-detects). Because *Legionella* spp. were detected in both cities (but never *L. pneumophila)*, a range of percentages for *pneumophila-like Legionella* (as a proportion of total *Legionella* spp.) was used to illustrate the potential range of risks involved (0.1 %, 1 %, 10 %, and 100 %); at least 20 *Legionella* spp. have been documented as human pathogens on the basis of their isolation from clinical material.^35^ In these simulations, *pneumophila-like Legionella* spp. were assumed to have the same dose-response characteristics as *L. pneumophila*, consistent with how pneumonia due to either *L. pneumophila* or *non-pneumophila Legionella* resemble each other clinically. ^36^ As an alternative supporting analysis, the concentration of *L. pneumophila* was estimated at the theoretical 95 % detection limit of the Legiolert assay (3 most probable number (MPN) in the total volume of water assayed).^37^ Because MAC was never detected while flushing showers in the St. Paul building, an upper limit was estimated based on the theoretical 95 % detection limit of the qPCR assay (i.e., 3 copies in the total volume of water analyzed by qPCR).^37^

Exponential dose-response models were used for both *Legionella* health endpoints based on a guinea pig model reported by Armstrong and Haas^38^, which has been validated against human outbreak data^39^ and with “uncertain parameter” values as reported in Hamilton et al.^32^ For MAC health endpoints, exponential dose-response models were used for pulmonary infection and cervical lymphadenitis (i.e., Tomioka model and Jorgensen 1 model, respectively), and an approximate beta-Poisson model for systemic infection (i.e., Yangco model).^31,40^

QMRA calculations were implemented in R software, using the package “mc2d” to conduct Monte Carlo simulations quantifying effects of uncertain inputs.^21,41^ Full equations and an example of R code are provided in the SI. All calculations were segregated by city (Minneapolis vs. St. Paul), and calculations involving aerosol inhalation were also segregated by shower head type. To assess the effect of flushing on risk, pathogen doses were calculated using either the concentrations from *t* = 0 min and 6 min, which represented the initial shower exposure after a stagnation period, or *t* = 15 min, 30 min, and 45 min for subsequent post-flushing exposure.

## Results

### Basic Water Quality Indicators

#### Total Chlorine

Total chlorine concentrations in Minneapolis pre-flush samples ranged from non-detect (i.e., <0.1 mg/L; 7/10 samples) to 1.0mg/L (Fig. S1). Total chlorine increased over time during cold water flushing and reached the respective building inlet concentrations (3.0 to 3.1 mg/L) within 30 min. Conversely, for hot water flushing, chlorine concentrations increased throughout the entire flushing period but reached only 22 to 91 % of building inlet values by the end of flushing. In the post-flushing period, chlorine concentrations declined rapidly with little remaining after 2 to 3 days in most cases (Table S3). Similar results were observed for the St. Paul building, except that the building inlet concentration (1.8 mg/L) was much lower than those in Minneapolis and concentrations in all post-flushing samples were negligible (≤0.1 mg/L).

In Minneapolis, chlorine decay within the shower supply lines was compared to that in clean glass bottles to assess the role of premise plumbing in chlorine consumption (Fig. S2). First-order chlorine decay rates (K) in the cold and hot water supply lines (1.04/day and 0.74/day, respectively) were much greater than the decay rate in clean glass bottles (0.05/day), suggesting a substantial chlorine demand associated with premise piping.

#### Temperature

The temperature of pre-flush water samples ranged from 19.7 to 30.4 °C (median: 22.6 °C) and either decreased over time for cold water flushes, approaching the building inlet temperatures, or increased over time to approximately 40 °C for most of the hot water flushes (Fig. S1, Table S4). Water temperature for one hot water flush in Minneapolis, however, did not increase substantially likely due to a faulty mixing valve. For the first hot water flush in St. Paul, the water became progressively colder because the water heating system initially was off. Water temperatures in the post-flushing period ranged from 24.2 to 29.1 ^°^C.

#### pH

The pre-flush pH ranged from 8.5 to 9.3 and tended to increase during cold water flushing up to the respective building inlet pH (9.2 to 9.5) and decrease slightly during hot water flushing (Fig. S1). The pH decreased over time in the post-flushing period for the cold-water flushed showers but was relatively stable for hot-water flushed showers (Table S5).

### Microbiological Indicators

#### Bacteria

Bacterial 16S rRNA genes were detected in all pre-flush samples at concentrations of 5.6 to 7.6 log_10_[copies/L] (Fig. 1). In Minneapolis buildings, concentrations decreased by 2 to 3 orders of magnitude within 6 min of flushing and stabilized at or near the respective building inlet concentrations (<method quantification limit (MQL) to 4.8 log_10_[copies/L]). In the St. Paul building, the decline during flushing was less apparent (Fig. 1) because the concentration in the inlet to the St. Paul building was relatively high (5.4 log_10_[copies/L]). Concentrations of bacterial 16S rRNA genes increased during the follow-up period. When all sites are considered together, pre-flush samples were significantly differentiated from samples collected following 30min of flushing (*p <* 0.001 via MLE) and from samples collected 2 to 4 days and then 6 to 7 days after flushing (*p <* 0.001 and *p =* 0.007, respectively, via MLE; Table S6).

**Figure 1.**
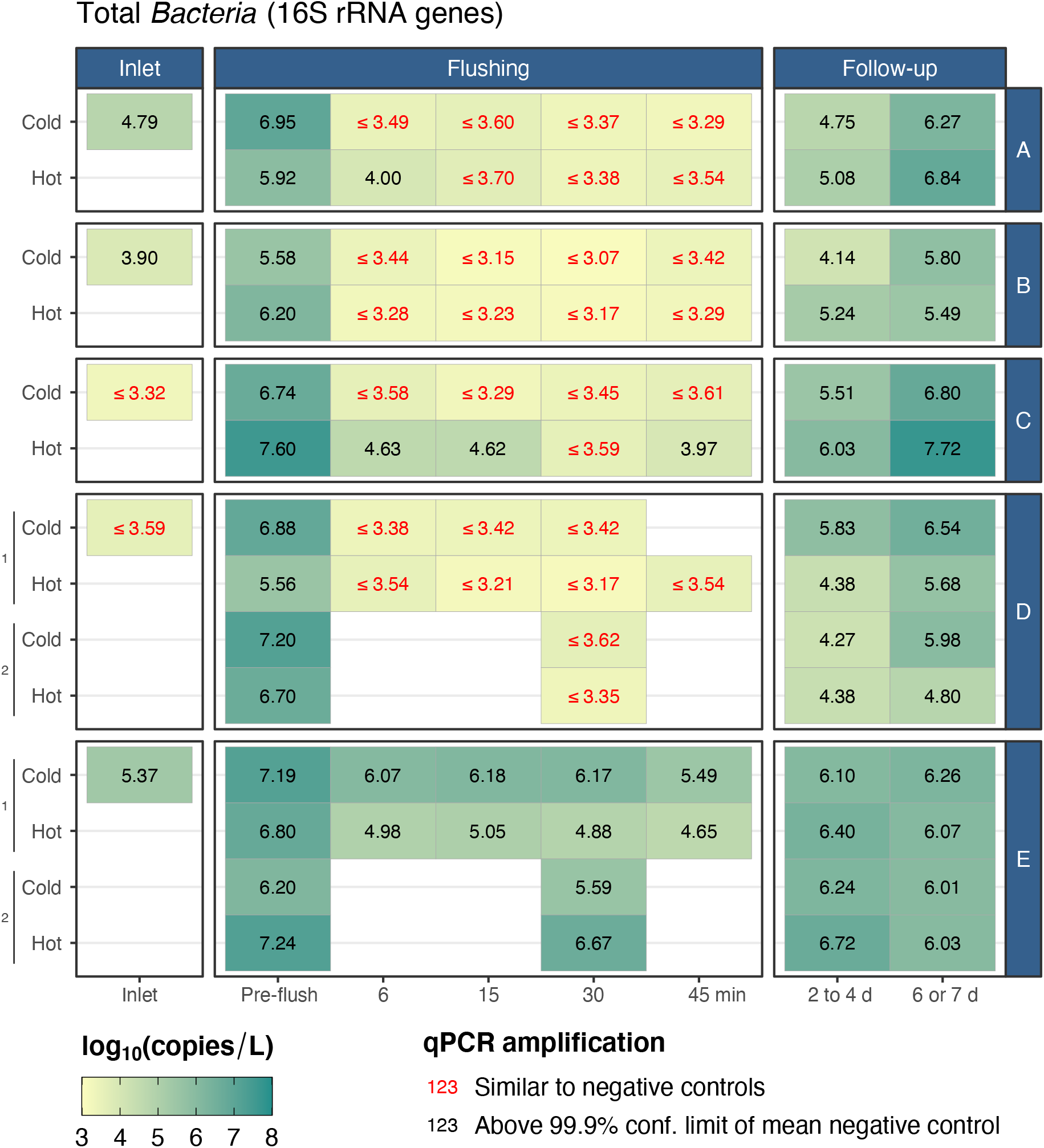
Heatmap of bacterial 16S rRNA genes (via qPCR) in Minneapolis (buildings A to D) and St. Paul (building E) premise water systems at the building inlet, during flushing of shower cold and hot water supplies in shower rooms, and during a follow-up monitoring period.

#### Legionellae

No culturable *L. pneumophila* were detected in 91 samples (260 Quanti-Trays total, including all technical replicates). All water samples were likewise negative for *mip* and *wzm* gene targets (L. *pneumophila* and *L. pneumophila* sg1).

Conversely, most pre-flush water samples in Minneapolis (8/10) and St. Paul (4/4) were positive for *ssrA (Legionella* spp.), with concentrations ranging from 4.4 to 6.2 log_10_[copies/L] (Fig. 2). The only two pre-flush samples that were negative for *Legionella* spp. were hot water samples. *Legionella* spp. were not detected at Minneapolis building inlets or in samples collected during flushing (i.e., 6 to 45 min) but were detected again during follow-up sampling in 20 % of showers after 2 to 4 days and 40 % of showers after 6 to 7 days.

**Figure 2.**
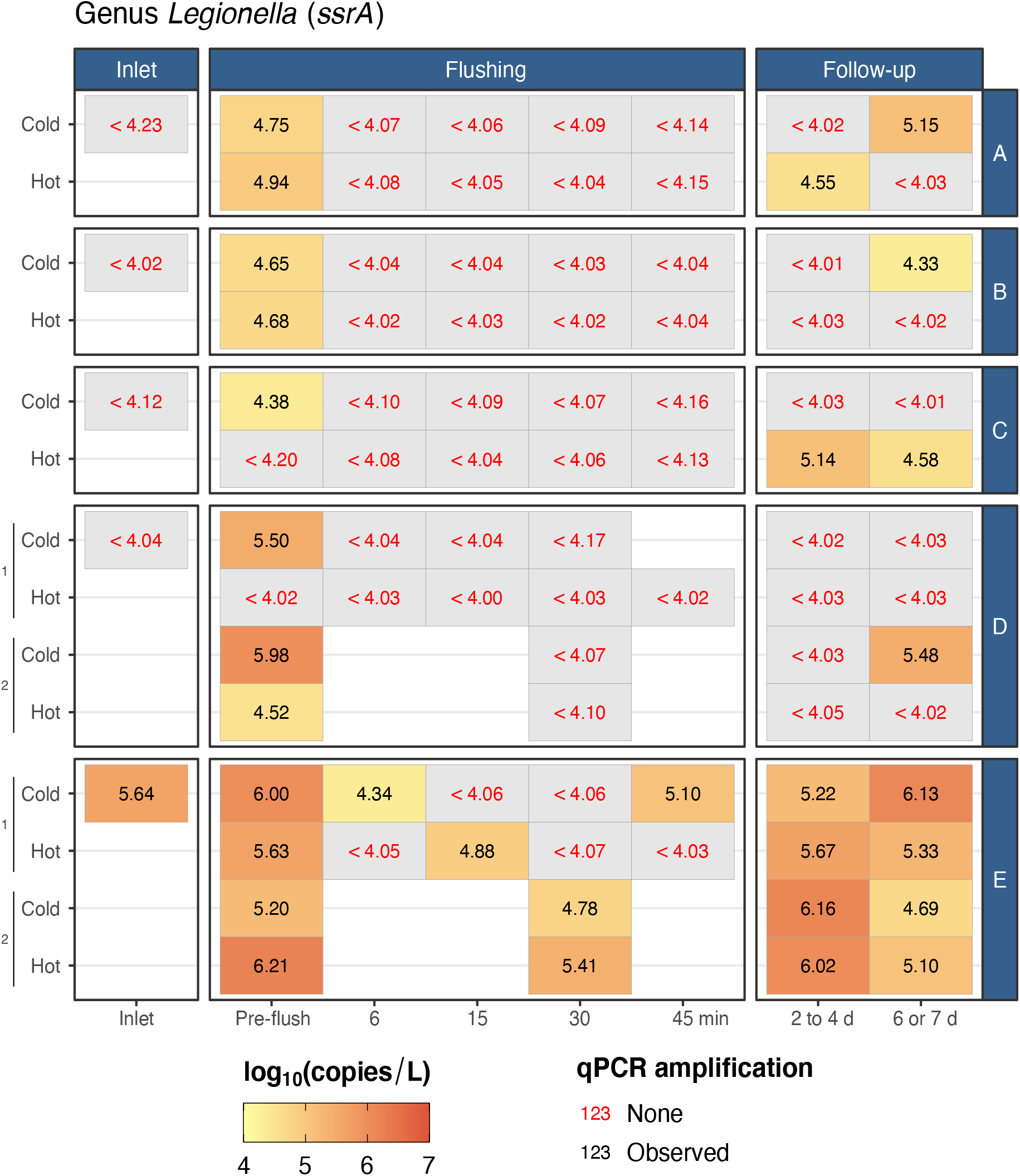
Heatmap of genus-wide Legionella ssrA genes (via qPCR) in Minneapolis (buildings A to D) and St: Paul (building E) premise water systems at the building inlet, during flushing of shower cold and hot water supplies in shower rooms, and during a follow-up monitoring period.

In St. Paul, *Legionella* spp. were detected at the inlet and in 5 of 10 water samples collected during flushing at 4.3 to 5.4 log_10_[copies/L], including 3 of 4 end-of-flush samples (30 or 45 min). The water heating system in building E was off during the first hot water flush (due to building disuse)—effectively another cold water flush, albeit from the hot water lines. The water heater was in operation, however, for the second flush. *Legionella* spp. were detected in all eight follow-up samples at concentrations ranging from 4.7 to 6.2 log_10_[copies/L]. Combining results from all buildings, *ssrA* gene concentrations were still less than pre-flush levels in the follow-up sampling after 6 to 7 days (*p =* 0.04 via MLE; Table S6).

#### Mycobacteria

In Minneapolis buildings, neither mycobacterial *atpE* nor MAC ITS were detected at building inlets but were detected in most pre-flush samples (9/10) at concentrations up to 7.6 log_10_[copies/L] and 5.4 log_10_[copies/L], respectively (Fig. 3). In the St. Paul building, *atpE* was detected at the inlet (3.8 log_10_[copies/L]) and in all four pre-flush samples (<3.1 to 5.8 log_10_[copies/L]), but MAC were never observed.

**Figure 3.**
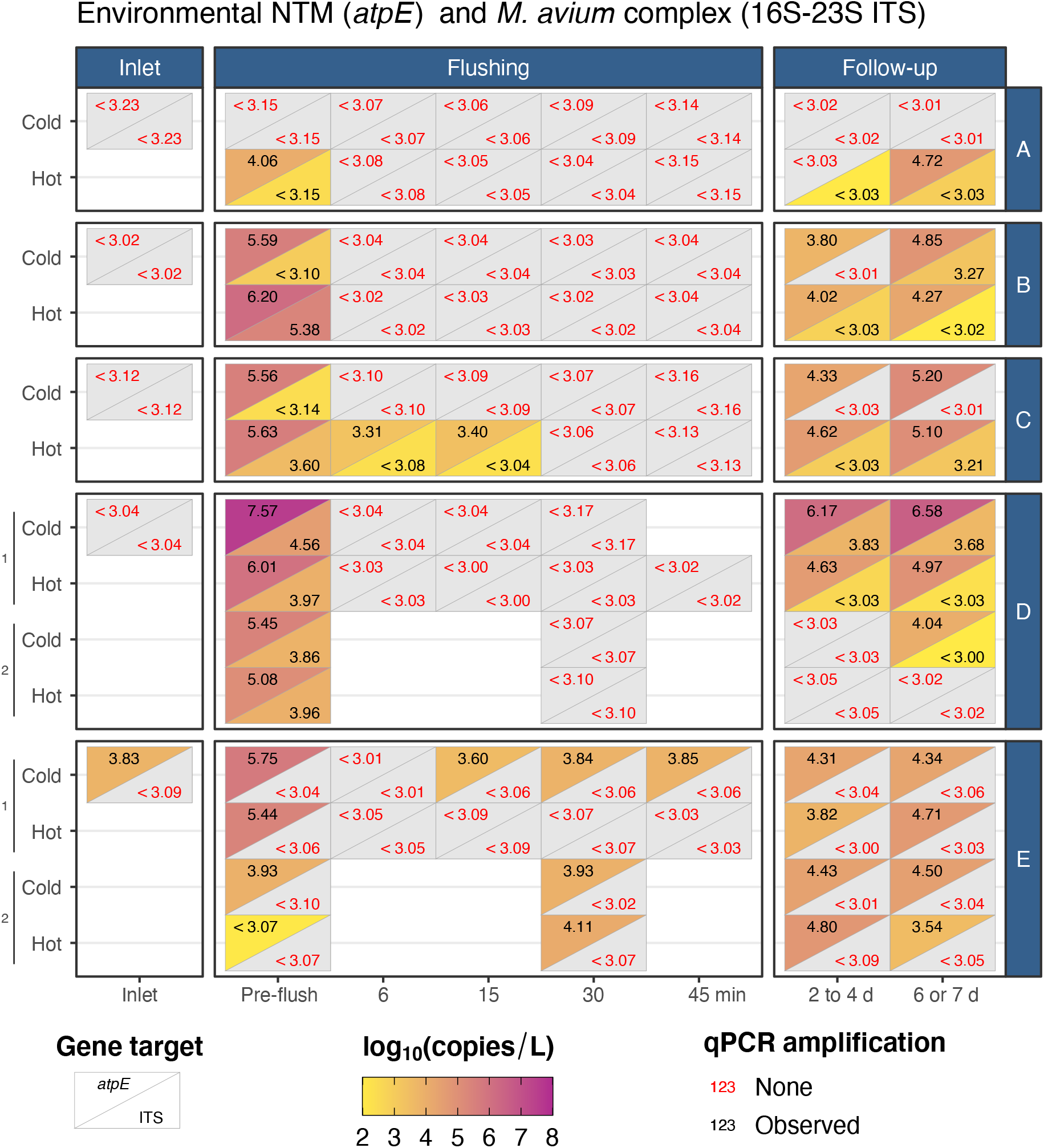
Heatmap of non-tuberculous mycobacteria (NTM) *atpE* genes and *Mycobacterium avium* complex 16S-23S internal transcribed spacer (ITS) region (via qPCR) in Minneapolis (buildings A to D) and St. Paul (building E) premise water systems at the building inlet, during flushing of shower cold and hot water supplies in shower rooms, and during a follow-up monitoring period.

Mycobacterial *atpE* concentrations—and in Minneapolis, MAC ITS concentrations—declined rapidly during flushing and were significantly reduced within 30min *(p =* 0.02 by MLE and *p =* 0.02 by Fong’s test for *atpE* and ITS, respectively; Tables S6 and S7). Mycobacterial *atpE* genes were detected again in 60% of Minneapolis and 100% of St. Paul samples collected 2 to 4 days after flushing. The concentrations of these genes, however, were still below pre-flush levels *(p =* 0.02 by MLE). The frequency of detection of Mycobacterial *atpE* genes increased to 80% in Minneapolis and 100% in St. Paul for samples collected 6 to 7 days after flushing, with concentrations still below pre-flush levels *(p =* 0.02 by MLE). In Minneapolis, MAC ITS concentrations were not significantly different from pre-flush levels at 6 to 7 days *(p =* 0.21 by Fong’s test).

#### Relationships Between Water Quality and Microbiological Indicators

After controlling for confounding effects of municipal water supply and building, total chlorine, temperature, and pH were fit to PCA biplots for cold and hot water (Fig. S3). Full statistical output is provided in the SI, including summary results of Spearman’s rank correlation tests (Tables S8 and S9). Total chlorine was strongly associated with gene marker concentrations (cold water: *r*^2^ = 0.57, *p =* 0.001, and hot water: *r*^2^ = 0.52, *p =* 0.001). Spearman’s rank tests indicated a strong negative correlation between gene markers and total chlorine, particularly in Minneapolis ******(*ρ* values = -0.84 to -0.49). Temperature likewise corresponded to gene marker concentrations (cold water: *r*^2^ = 0.57, *p =* 0.001, and hot water: *r*^2^ = 0.21, *p =* 0.008). Spearman’s rank tests indicated positive correlation with temperature (ρ up to 0.80) for cold water—and hot water at building E, which had not been heated in one case—and negative correlation for hot water in Minneapolis *****(ρ down to -0.59). Water pH was weakly associated with gene marker concentrations in cold water (*r*^2^ = 0.14, *p =* 0.03) but not hot (*r*^2^ = 0.06, *p =* 0.3). Spearman’s rank tests indicated a negative correlation between gene markers and pH, but only in Minneapolis cold water (****ρ = -0.55 to -0.66).

### Human Health Risks

#### Legionellae

*L. pneumophila* risk estimates were typically lower than a daily equivalent of the U.S. Environmental Protection Agency (EPA) 10^-4^ annual acceptable risk threshold (10^-4^/365 = 3 × 10^-7^ infections per day; Fig. 4, with full results detailed in Tables S10 and S11). The median risk of subclinical infection (per showering exposure) ranged from 10^-8^ to 10^-7^, while the median risk of clinical severity infection was between 10^-11^ and 10^-10^ per exposure. Note that all *L. pneumophila* risk estimates presented here are likely to be upper limits on true risk, as *L. pneumophila* was never detected in Minneapolis or St. Paul.

**Figure 4.**
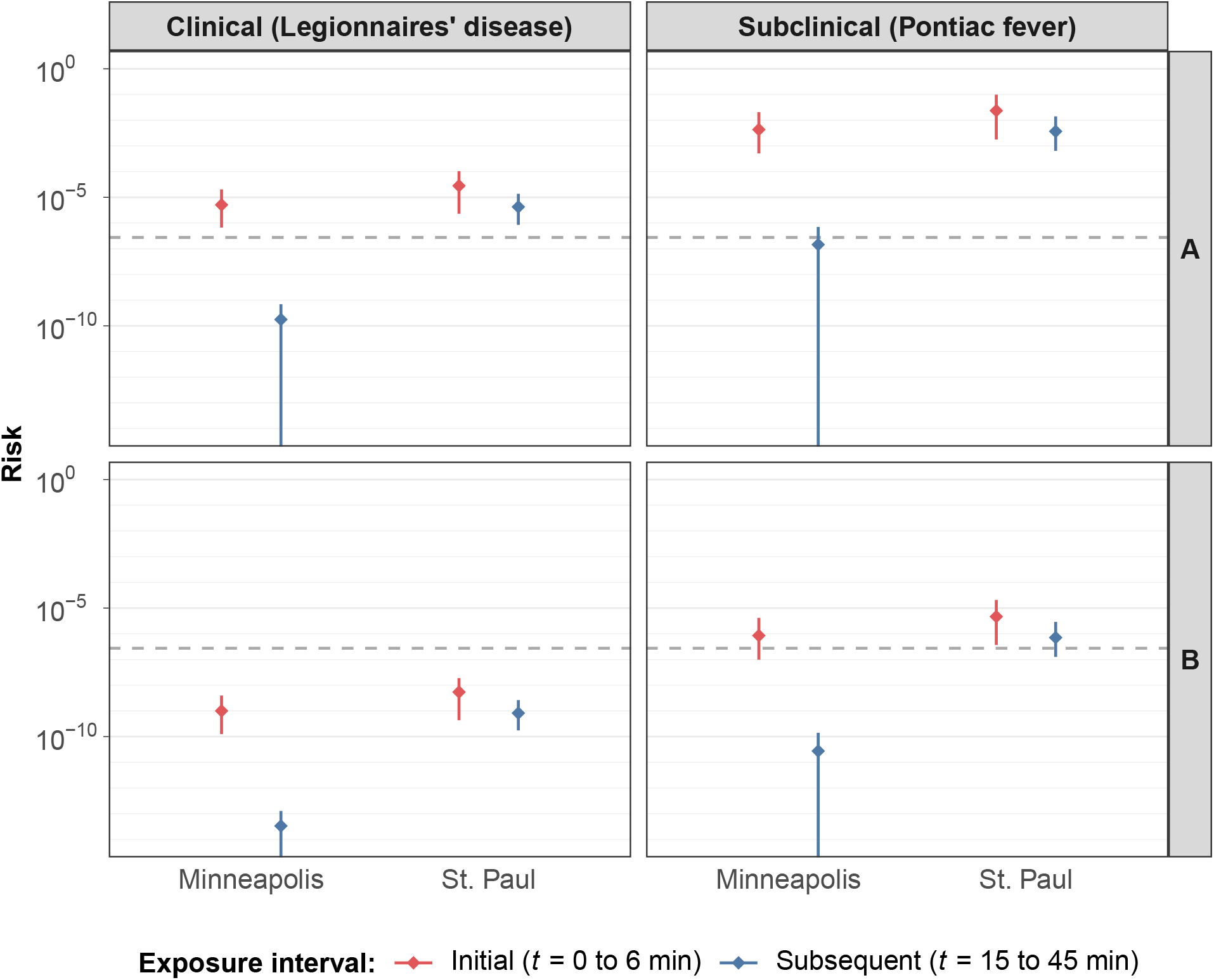
Per-exposure risk (median ± 95% confidence interval via Monte Carlo simulation) of clinical and subclinical legionellosis in the Minneapolis and St. Paul university shower rooms, for the initial shower after stagnation versus the subsequent post-flushing interval. Scenario A (top) assumes 100% *pneumophila-like Legionella* spp. using a conventional shower head, while Scenario B (bottom) assumes 0.1 % *pneumophila-like Legionella* spp. using a water-efficient shower head. Dashed line indicates a daily acceptable risk threshold.

Initial exposure risk estimates for *Legionella* spp.-induced health impacts varied widely and were linearly related to the percentage of *Legionella* spp. assumed to be *pneumophila-like* (Tables S10 and S11). Risks of subclinical infection (per exposure) were as low as 10^-6^ if only 0.1 % of *Legionella* spp. were assumed to be *pneumophila-like*, whereas they exceeded 10^-2^ if 100 % of *Legionella* spp. were assumed to be *pneumophila-like*. Risks of clinical severity infection illustrated a similar range, though were about 1000-fold lower than the corresponding risks of subclinical infection, which corresponded with the differences in dose-response models for these two health endpoints. Finally, all risk estimates for *pneumophila-like Legionella* were 10-fold or more greater than those for *L*. *pneumophila* assuming *L*. *pneumophila* was present at the Legiolert lower limit of detection. Thus, if no *Legionella* spp. were *pneumophila-like*, then overall risks from exposure to legionellae are substantially overestimated.

Regardless of differences between the *L*. *pneumophila* and *pneumophila*-like assumptions, differences between shower head type were consistent. Risk for conventional shower heads was always about 5 times higher than risk for efficient shower heads, which corresponded to the assumed model parameter differences for these two fixture types. This can likely be attributed to the lower flow rates of efficient shower heads, as well as to different aerosol size distributions between the two.

Differences in initial exposure risk estimates between cities were also generally consistent between *L*. *pneumophila* and *pneumophila*-like *Legionella*. For *L*. *pneumophila*, risk was higher in the St. Paul building compared to that in the Minneapolis buildings by a factor of 3, and for *pneumophila-like Legionella*, risk estimates were higher in St. Paul by a factor of nearly 8. In the former case, the higher estimated risk in St. Paul resulted from the smaller total volume of water sampled (14 flushing period samples in St. Paul compared to 42 in Minneapolis). In the latter case, higher risk in St. Paul is attributed to the relatively high concentrations of *Legionella* spp. after 6 min of flushing (Fig. 2). There were substantial differences in the benefits of flushing with respect to infection risk for buildings in the two cities. Flushing reduced the risk by more than 4 orders of magnitude in the Minneapolis buildings compared to only one order of magnitude in the St. Paul building. This is attributed to the relatively high concentrations of *Legionella* spp. throughout the flushing period likely resulting from the relatively high concentrations in the St. Paul building inlet water.

#### MAC

In general, MAC risk estimates were relatively low (Fig. 5, with full results detailed in Tables S12 and S13). The risk for pulmonary infections from inhalation exposures were all 2.4 x 10^-9^ or lower, regardless of whether the samples were from the initial exposure after stagnation or subsequent flushing period. Median risk estimates for illness from ingestion exposures were in the range of 2.7 x 10^-11^ to 1.5 x 10^-6^, with the higher value for systemic infection from pre-flush samples in Minneapolis. The difference in risk for the two exposure routes is largely due to differences in assumed water exposure (4 to 20 μL of aerosolized water versus 0.1 to 2.0 mL of accidentally ingested water). Between the two health endpoints relevant for ingestion exposure, risk estimates for systemic infection were about 10-fold higher than those for cervical lymphadenitis, which corresponded to differences in the dose-response models for these two endpoints. Between cities, risk estimates for the buildings in Minneapolis were about 700-fold higher than for the building in St. Paul, which corresponded to differences in water concentrations between cities. In particular, MAC were never detected in St. Paul, so as noted earlier, MAC risk estimates in St. Paul were based on the theoretical detection limit of the qPCR assay. Note also that all MAC risk estimates are likely to be conservatively high, because the dose harmonization factor was assumed to be 1, while in reality it is probably higher (e.g., it was assumed to be 10 to 30 for legionellae).

**Figure 5.**
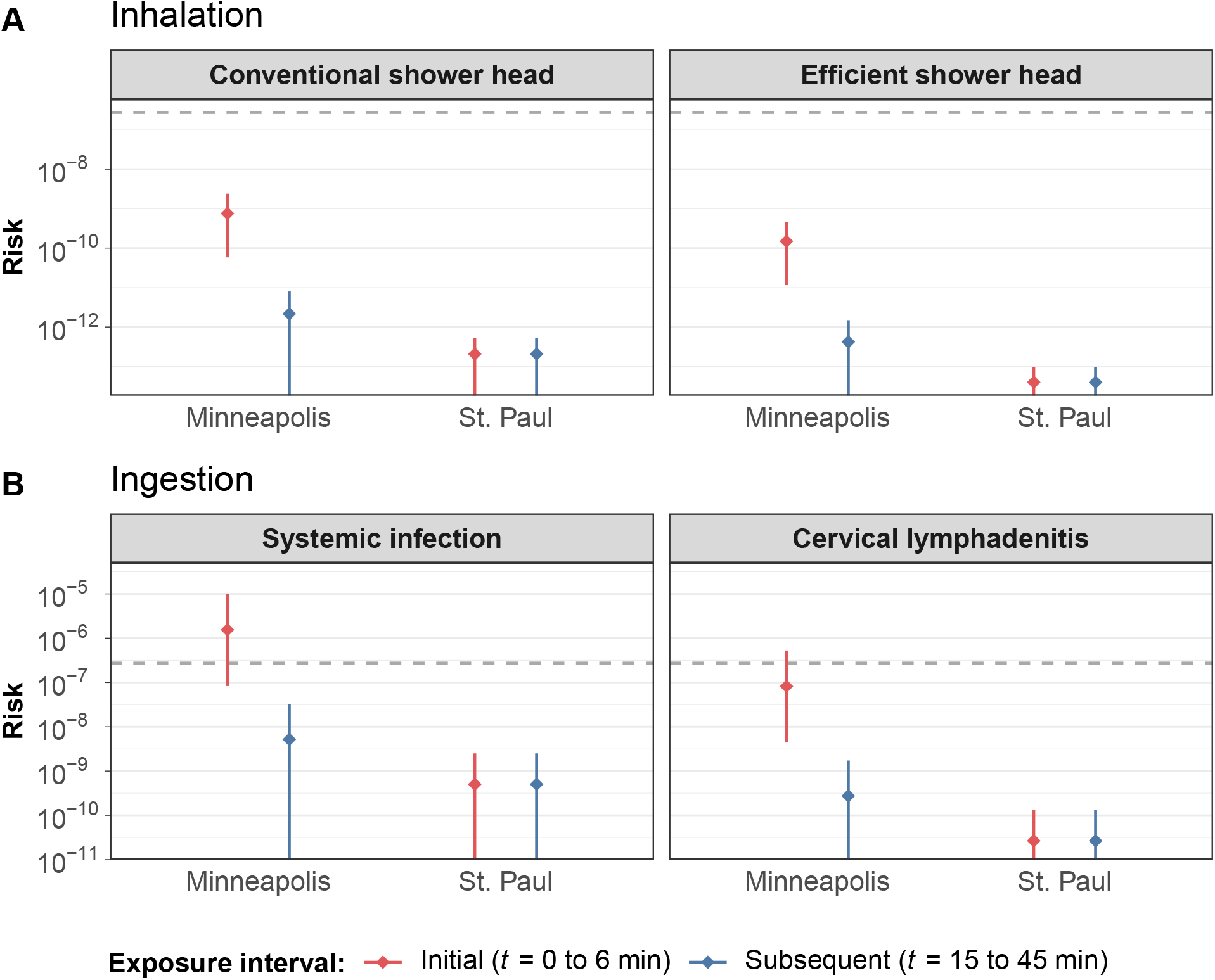
Per-exposure risk (median ± 95% confidence interval via Monte Carlo simulation) for the initial shower after stagnation versus the subsequent post-flushing interval in the Minneapolis and St. Paul university shower rooms, resulting in either (**A**) pulmonary infection via *Mycobacterium avium* complex (MAC) inhalation using either conventional or water-efficient shower heads, and (**B**) systemic infection or cervical lymphadenitis via ingestion of MAC. Dashed line indicates a daily acceptable risk threshold.

### Discussion

Pre-flush samples from idle showers serviced by two independent chloraminated distribution networks were uniformly negative for L. *pneumophila* using both cultivation-based and cultivation-independent methods. The lack of *L. pneumophila* is a significant finding from a public health perspective, because *L*. *pneumophila* are well-known to cause the potentially fatal pneumonia called Legionnaires’ disease and the less serious flu-like illness called Pontiac fever. These results are consistent with previous reports that premises receiving chloraminated drinking water tend to have lower levels of legionellae compared to chlorinated drinking water.^10,12^ Although limited to two systems, the results from this study expand on prior work by suggesting that chloramine affords protection from *L*. *pneumophila* colonization of premise plumbing and subsequent proliferation during extended water stagnation.

Despite this promising finding, *Legionella* spp. were still observed in pre-flush samples by qPCR targeting ssrA, indicating that some *Legionella* spp. survive in these chloraminated premise plumbing environments. Additionally, the re-occurrence of *Legionella* spp. during follow-up monitoring suggests these organisms were present in premise plumbing biofilms or in the water at the end of flushing at concentrations less than the detection limit of our assay. Bédard et al.^42^ attributed the rapid increase in culturable cell density after short stagnation times (1 day) to bacterial detachment from the biofilm as opposed to cell growth or restoration of culturability. As our post-flushing samples were obtained with up to 7 days of stagnation, cell growth may also be a factor. To assess the risk posed by the legionellae in these plumbing systems, additional work would be needed to determine the strains present and their proclivity to cause human disease.

Neither NTM nor MAC were detected in the building inlets in Minneapolis but were present in most pre- and post-flushing samples. In contrast, NTM were detected in the inlet, during flushing, and in the follow-up sampling of the St. Paul building, although MAC were never observed. The rapid rebound of NTM, MAC, or both in the post-flushing samples in this study is consistent with their presence in premise plumbing biofilms. It is not surprising to find NTM in these premise plumbing systems because NTM are predominant members of biofilm communities in chloraminated distribution networks^17,18^ and may also reside in premise plumbing biofilms.^43^ The proliferation of NTM in these environments is attributed to their well-known resistance to disinfectants and other antimicrobial agents.^44^ Despite our previous observations of abundant NTM in two chloraminated systems, MAC were not detected in any water or biofilm samples collected directly from the distribution mains of those systems.^17,18^ A key factor that may explain this apparent discrepancy is the low to negligible total chlorine concentrations in the premise plumbing versus relatively high concentrations in distribution mains (approximately 3.5mg/L). Haig et al.^19^ reported a correlation between *M. avium* abundance in water samples and water age, which accounted for distribution network travel time and stagnation time in premise plumbing. Despite correlating with water age, surprisingly, chloramine concentration—averaging 2.04 mg/L—was not identified as a statistically significant parameter correlating to *M. avium* abundance.^19^ The presence of MAC in the Minneapolis showers poses some, albeit minor, risk given the potential for MAC to cause fatal pneumonia in people with compromised immune systems or chronic lung conditions. ^5^

Few QMRAs for opportunistic pathogens have examined risks associated with indoor use of distributed potable water based on site-specific measurements. Instead, prior work has largely focused on reclaimed water use, greywater reuse, and/or roof-harvested rainwater use.^31,45,46^ Additionally, Hamilton et al. ^32^ worked backwards from target risk values to derive critical concentrations informing monitoring efforts for L. *pneumophila* exposures associated with indoor use of potable water. Thus, the current QMRA is novel in its reliance on site-specific empirical measurements of *Legionella* spp. from distributed water systems, and we are aware of only one other comparable study in the peer-reviewed literature.^47^ Assuming a conversion factor of 0.97 disability-adjusted life years (DALYs) per subclinical infection^32^ and 365 daily exposures per year, the Sharaby et al.^47^ risk estimates correspond to probabilities of subclinical infection equal to approximately 2 x 10^-6^ and 7 x 10^-6^ per person per daily exposure for faucet use and showering, respectively. These risk estimates are consistent with the low end of the range for pre-flush samples reported in the current study, when 0.1 % of *Legionella* spp. are assumed to be *pneumophila-like*.

Considering previous studies investigating other types of source water and exposures, the risk estimates for L. *pneumophila* and *pneumophila-like Legionella* spp. from the present study are generally comparable. Blanky et al.^45^ estimated annual probabilities of subclinical L. *pneumophila* infection between 10^-8^ and 10^-5^ for garden irrigation with greywater and between 10^-6^ and 10^-4^ for toilet flushing with greywater. Meanwhile, Hamilton et al.^31^ estimated annual probabilities of subclinical L. *pneumophila* infection between approximately 10^-6^ and 10^-2^ for a number of exposure scenarios involving roof-harvested rainwater. In addition, Hamilton et al.^46^ estimated annual probabilities of subclinical L. *pneumophila* infection between 10^-6^ and 10^-2^ for toilet flushing with reclaimed wastewater. When converted to daily risk (assuming 365 daily exposures per year for any given activity), the majority of these risk estimates fall well below 10^-5^ subclinical infections per person per daily exposure, which correspond to the low to mid-range risk estimates for the current study.

Furthermore, when compared to the EPA annual acceptable risk threshold of 10^-4^ infections per person per year (which equates to approximately 3 x 10^-7^ infections per person per day), most *L*. *pneumophila* risk estimates were also relatively low, with the exception of subclinical infection risks for conventional shower fixtures in St. Paul (2.7 x 10^-7^). Additionally, pre-flush risks of subclinical infection for *pneumophila-like Legionella* spp. all exceed the daily equivalent of the EPA annual threshold but could be substantially reduced by flushing, especially in the Minneapolis buildings. It should be emphasized, however, that the true percentage of *pneumophila-like Legionella* spp. in the currently study is unknown, and the assumed values were selected arbitrarily to illustrate trends among a range of possible scenarios. Future work quantifying the proportion of highly infectious (i.e., *pneumophila-like) Legionella* spp. in tap water or premise plumbing biofilms, or both, would therefore be particularly valuable for refining these risk estimates.

Previous QMRA studies for MAC are extremely limited. Rice et al. ^48^ considered exposure of individuals with acquired immune deficiency syndrome (AIDS) to MAC via distributed drinking water but only estimated probabilities of exposure rather than probabilities of infection or illness because dose-response models for MAC were unavailable at the time of their study. Hamilton et al.^31^ performed a risk assessment for roof-harvested rainwater and considered a wide variety of exposure routes for MAC, including 9 ingestion routes and 5 inhalation routes. Ingestion-associated risk estimates, including from intentional consumption of drinking water, were generally higher than inhalation-associated risk estimates for MAC (10^-7^ to 1 versus 10^-10^ to 10^-4^, respectively). MAC risk estimates from the present study, with median probabilities of infection (per single shower exposure) between approximately 10^-11^ and 10^-6^, are on the lower end of values reported by Hamilton et al.^31^

It is important to note that our QMRA analysis considered showering exposures only and excluded exposures from other common indoor water uses such as toilet flushing and the use of faucets. This choice was justified based on previous work demonstrating that showering presents the highest risk compared to other indoor water uses.^32^ In addition, the current analysis has focused on per exposure (i.e., per showering event) risk estimates rather than annual risk estimates because the concern was about exposure to stagnant water following an extended period of building disuse, which is unlikely to occur repeatedly throughout an entire year.

There were distinct differences in total chlorine and bacterial gene target concentrations for the building inlets in Minneapolis versus the one in St. Paul despite the fact that the water for both systems is primarily sourced from the Mississippi River, treated similarly, and distributed with a similar residual chloramine concentration. The likely explanation for the observed water quality differences is that the water in St. Paul was older, due to the presence of a university-owned water storage tower combined with low water use on the campus due to the COVID-19 pandemic. The differences in building inlet water qualities also translated into substantial differences in how the showers on each campus responded to flushing and and the resulting post-flushing health risks from *Legionella* spp. exposure.

The total chlorine residual in all buildings was unstable, rapidly decaying during post-flushing stagnation. Decay rates were 14 to 19 times greater in the premise plumbing compared to clean glass bottles, which is consistent with a report by Rhoads et al.^49^, who observed high chloramine decay rates in green building water systems. The microbiological results suggest that biofilms are present in the plumbing, and biofilms are known to exert a chloramine demand, especially nitrifying biofilms.^50^ The observed pH decrease during post-flushing stagnation is consistent with ammonia-oxidizing microorganism activity. Plumbing components or substances associated with those components may also contribute to chlorine decay. For example, copper ions from actively corroding copper pipes can exert a chlorine demand.^51^ The negative correlation between total chlorine and marker genes is consistent with the well-known effects of residual disinfectant on bacterial growth. All bacterial marker gene concentrations rapidly increased during the post-flushing period and after 6 to 7 days approached those in the pre-flush water that had stagnated for about 2 months. This further supports the need for frequent flushing to maintain a residual in plumbing, in addition to adequate cold or hot water temperatures, as control measures against bacterial growth during a shutdown. Guidance for recommended residual levels is ≥0.2 mg/L for free chlorine^52^ but less formally defined for chloramine, though total chlorine as low as 0.1 mg/L may still afford some protection against *Legionella*.^16^

Despite the overall low health risks from opportunistic pathogen exposure observed in this work, the results have important implications for flushing practice. First, flushing showers with chloraminated cold water or hot water may be able to reduce *Legionella* spp., NTM, and MAC concentrations to below detection limits in as little as 6 min. Although temperature was significantly associated with bacterial gene markers, positively in cold water and negatively in hot water, this was likely due to the arrival of fresh water containing a residual disinfectant and lower bacteria concentrations. Nevertheless, when the building inlet water contains microorganisms of potential health concern like legionellae or NTM, preliminary flushing of local water mains and water turnover in nearby storage tanks may be necessary. Finally, given that water quality declines rapidly during stagnation, further consideration should be given to flushing periodically during shutdowns, or at a minimum, flushing within 2 or 3 days of building re-occupancy. Certainly, every building is unique and flushing requirements may be different for other locations.

## Data Availability

Data are provided in the manuscript or supporting information file.

## Acknowledgement

The authors thank Scott Bernardson, Kirk Hall, and Tony Gutterman from UMN Facilities Management for their assistance with building access and sampling. The authors also thank personnel at the Minnesota Department of Health (MDH), especially Anita Anderson, Kim Larsen, and Alex Bartley, for guidance in the development of a sampling and flushing plan. Finally, the authors thank MDH for financial support of this research.

## Supporting Information Available

Supplemental experimental materials and methods, including sample collection processing, realtime qPCR, data analysis and statistics (with example R software code), and QMRA equations (with example code); supplemental results, including relationships between water quality and microbiological indicators (with R software hypothesis testing console output); supplemental tables summarizing building sites, sample collection dates, total chlorine observations, water temperature observations, water pH observations, paired MLE contrasts, Fong’s modified sign test contrasts, Spearman’s rank correlations for Minneapolis and St. Paul, all QMRA estimates for simulated *Legionella* exposure via inhalation and MAC exposure via inhalation and ingestion in both Minneapolis and St. Paul, measured and predicted hydraulic characteristics during flushing, and qPCR calibration curves; supplemental figures showing water quality during flushing, chloramine decay and decay rates, qPCR of bacterial 16S rRNA genes, and PCA biplots of marker genes.

## SI Figures & Tables

This page is for LaTeX cross-referencing to the Supporting Information in this draft version of the manuscript only.

**Figure S1**. Water quality during flushing

**Figure S2**. Chloramine decay in Minneapolis water

**Figure S3**. PCA biplot of ranked marker gene concentrations

**Table S1. Building site descriptions**

**Table S2. Sample collection dates**

**Table S3. Total chlorine summary**

**Table S4. Water temperature summary**

**Table S5. Water pH summary**

**Table S6. Paired MLE contrasts**

**Table S7. Fong’s modified sign test contrasts**

**Table S8. Spearman’s rank correlations, Minneapolis**

**Table S9. Spearman’s rank correlations, St. Paul**

**Table S10. Full QMRA results for *Legionella* exposure (Minneapolis)**

**Table S11. Full QMRA results for *Legionella* exposure (St. Paul)**

**Table S12. Full QMRA results for MAC exposure via inhalation**

**Table S13. Full QMRA results for MAC exposure via ingestion**

**Table S14. Measured and predicted hydraulic characteristics**

**Table S15. Real-time qPCR summaryGraphical TOC Entry**

## Graphical TOC Entry

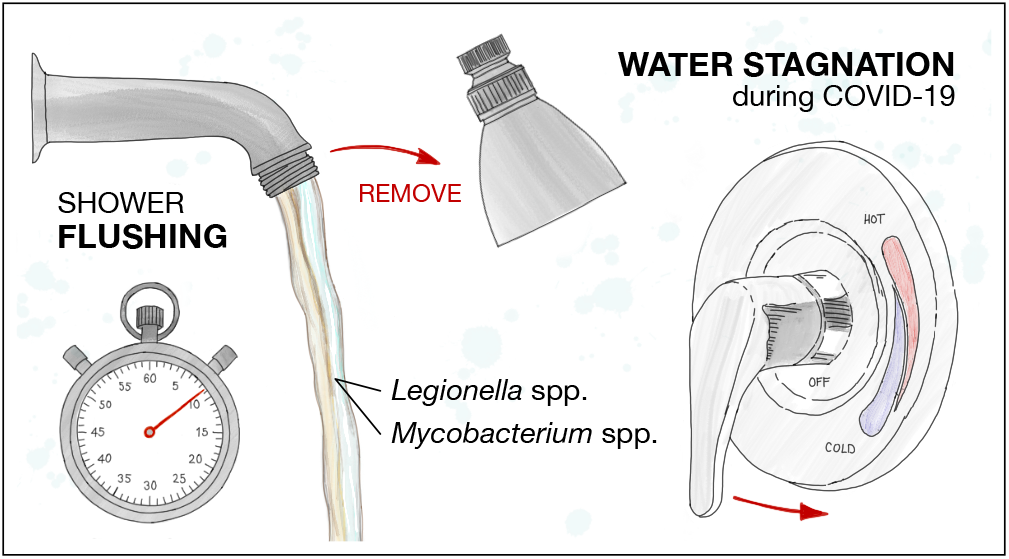

## Notes

### Competing Interest Statement

The authors have declared no competing interest.

### Author Declarations

No IRB oversight needed as no sampling or testing of humans was done.

